# Insomnia symptom prevalence in England: A comparison of self-reported data and primary care records in the UK Biobank

**DOI:** 10.1101/2023.09.07.23295191

**Authors:** Melanie A de Lange, Rebecca C Richmond, Sophie V Eastwood, Neil M Davies

## Abstract

**Purpose:** We aimed to use a large dataset to compare self-reported and primary care measures of insomnia symptom prevalence in England and establish whether they identify participants with similar characteristics.

**Methods:** We analysed data from 163,748 UK Biobank participants in England (aged 38-71 at baseline) with linked primary care electronic health records. We compared the percentage of those self-reporting ‘usually’ having insomnia symptoms at UK Biobank baseline assessment (2006-2010) to those with a Read code for insomnia symptoms in their primary care records prior to baseline. We stratified prevalence in both groups by sociodemographic, lifestyle, sleep and health characteristics.

**Results:** We found that 29% of the sample self-reported having insomnia symptoms, whilst only 6% had a Read code for insomnia symptoms in their primary care records. Only 10% of self-reported cases had an insomnia symptom Read code, whilst 49% of primary care cases self-reported having insomnia symptoms. In both primary care and self-reported data, prevalence of insomnia symptom cases was highest in females, older participants and those with the lowest household incomes.

However, whilst snorers and risk takers were more likely to be a primary care case, they were less likely to self-report insomnia symptoms than non-snorers and non-risk takers.

**Conclusions:** Only a small proportion of individuals experiencing insomnia symptoms present to primary care. However, the sociodemographic characteristics of people attending primary care with insomnia were consistent with those with self-reported insomnia, thus primary care records are a valuable data source for studying risk factors for insomnia.

**Key Points:**

- Around a third of the general population is thought to suffer from insomnia symptoms, but estimates are based on small samples and rely on people self-reporting their symptoms.
- Electronic health records (EHRs) offer a more objective means of measuring insomnia prevalence, but small-scale studies suggest they only capture a small proportion of insomnia cases. It is therefore unclear how useful EHRs are in measuring the prevalence of insomnia.
- In a sample of over 160,000 UK Biobank participants in England we found that 29% of participants self-reported having insomnia symptoms, whilst only 6% had a Read code for insomnia symptoms in their primary care records.
- Characteristics of people attending primary care with insomnia symptoms are similar to those self-reporting insomnia symptoms, suggesting EHRs offer a valuable data source for studying risk factors for insomnia.

**Plain Language Summary:** Around a third of the general population is thought to suffer from insomnia symptoms, but estimates are based on the responses of a small number of people and rely on them reporting their own symptoms. People’s medical records offer a more objective way of finding out how many people have insomnia, but only capture people who go to their doctor for help. In this study we compared 160,000 people’s answers to a question on insomnia symptoms to their primary care records. We found that 29% of people reported insomnia symptoms, whereas only 6% had insomnia symptoms recorded in their medical records. However, the characteristics of those reporting insomnia and those with insomnia in their medical records were similar. This means that although medical records only capture a small proportion of those suffering from insomnia, they do still provide useful information for researchers studying risk factors for insomnia.

## 1. INTRODUCTION

Insomnia is a distressing, yet common, condition which has extensive consequences for population health^1^. It has been associated with a variety of health problems including depression^2, 3^, substance use^4, 5^, dementia^6^, diabetes^7^ and cardiovascular disease^8, 9^. In addition, insomnia has been associated with lower productivity^10^ and higher absenteeism in the workplace^11, 12^, higher accidents rates^11, 13^, greater healthcare utilisation^11, 12^ and reduced quality of life^12, 14^.

Estimates of insomnia prevalence differ depending on the definition used. A review of 50 studies from different countries found that around a third of the general population have insomnia symptoms, 9-15% suffer from daytime consequences of insomnia, 8–18% are dissatisfied with their sleep and 6% meet the criteria for an insomnia diagnosis^15^. However, most previous studies estimating insomnia prevalence rely on participants self-reporting their symptoms and diagnoses through questionnaires or telephone interviews. Responses may therefore be subject to recall bias^16^. Many surveys have used small sample sizes consequently limiting their precision^17^. Selection bias could also be an issue in these studies if survey non-response was also related to insomnia prevalence^18^.

In recent decades there has been a rapid growth in the use of electronic health records (EHRs) in population health research^19^. EHRs potentially offer larger sample sizes, rich longitudinal data, lower risk of recall bias and, in countries such as the UK (where 98% of the population is registered with a primary care doctor and consultations are free of charge^20^), can reduce selection bias^19^. However, to date, EHR research on insomnia prevalence is limited. One US study of 15 family practices (n=7,928) found that 9.4% of primary care patients had an insomnia diagnosis, 7.4% had been prescribed an insomnia-related medication and 3.9% had both a diagnosis and prescription. Diagnoses and prescriptions were greater in women than men, and increased with age^21^. Another study found that 15% of a sample of 440,000 US Veterans had a prescription for an insomnia medication, whilst 6% had a diagnosis for insomnia^22^.

By definition EHRs only capture events where a patient visits a health care professional. As a result, mild or temporary conditions may be missed^22^. This is particularly likely to be the case for insomnia where only around a third of those self-reporting insomnia symptoms also self-report seeking help from a healthcare provider for them^23-25^. Only a few studies have explored this gap in data capture by comparing people’s self-reported insomnia symptoms to their actual medical records. One study in Majorca found that of patients who met the criteria for an insomnia diagnosis during a telephone survey (n=99), only 40% had a consultation for insomnia and only 12% had an insomnia diagnosis in their medical record. Another study of 5 UK GP practices (n=327) found that whilst 34% of patients self-reported insomnia symptoms, only 19% of this group had a primary care consultation for insomnia or a mood problem in the following 12 months and 30% had a consultation or prescription for insomnia/ mood problem^26^. These studies suggest that EHRs are only picking up a small proportion of people experiencing insomnia symptoms. However, the generalisability of this research is limited due to small sample sizes and their results are yet to be replicated in larger studies. It is therefore not clear how useful EHRs are in measuring the prevalence of insomnia.

Using UK Biobank data, which combines self-reported measures of insomnia with linked primary care records for over 160,000 people in England, this study aimed to compare self-reported and primary care measured insomnia symptom prevalence. We also aimed to establish whether self-report and primary care insomnia data identify participants with similar characteristics in order to evaluate the value of EHRs in measuring insomnia prevalence.

## 2. METHODS

### 2.1 Study Population

The UK Biobank is a population-based cohort study of around 500,000 adults who were aged 39-69 when recruited from across the UK between 2006 and 2010 (participation rate: 5.5%)^27^. It contains comprehensive questionnaire data, as well as physical measurements and biological samples^27^. Linked primary care data is also available for around 45% of UK Biobank participants^28^.

Of the 502,387 participants, we removed 338,197 who did not have linked primary care registration data provided by TPP (an England-only dataset). We also removed 6 participants who did not have a registration date in their primary care registration data and 436 participants without data on self-reported insomnia. This gave us a sample of 163,748 participants (see Figure 1).

**FIGURE 1.**
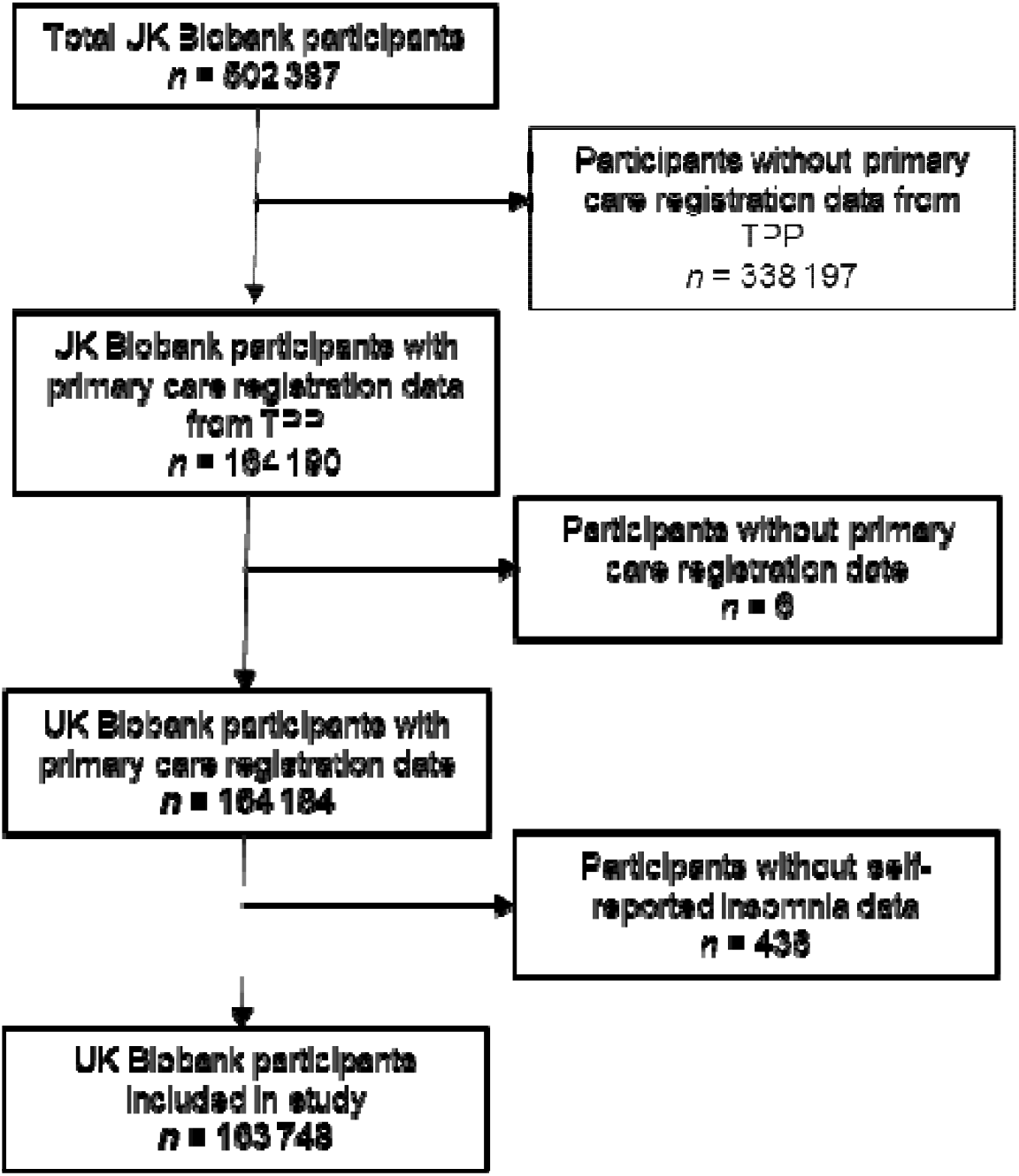
Participant flow diagram

### 2.2 Insomnia symptom case identification

#### Self-reported insomnia symptoms cases

Participants were asked in a UK Biobank touchscreen questionnaire: “Do you have trouble falling asleep at night or do you wake up in the middle of the night?”. If they pressed the help button they were told: “If this varies a lot, answer this question in relation to the last 4 weeks”. Options for participants’ answers were: “Never/rarely”, “Sometimes”, “Usually” and “Prefer not to answer”. We coded “Prefer not to answer” as missing. Participants were counted as a self-reported insomnia symptoms case if they answered “Usually”.

#### Primary care insomnia symptoms cases

Participants were designated as having primary care insomnia symptoms if they had a Read code for insomnia symptoms on or prior to UK Biobank baseline assessment (2006-2010). Insomnia Read codes occurring after baseline were excluded from our analysis in order to be consistent with the self-report question, which asked about existing symptoms. Read codes for events outside the date of an individual’s registration period at their GP practice (or practices) were excluded as coverage of symptoms during these periods may be unreliable. Sensitivity analyses defined insomnia by Read code and concomitant hypnotic use (see below).

#### Codelist creation

To create our list of Read codes for insomnia symptoms we searched the UK Biobank Read CTV3 code lookup table for codes containing the following strings: *insomn* *sleep* or *wak*. This gave us an initial list of 385 insomnia Read codes. This list was then refined by primary care clinician review. We included codes involving physical (organic) causes of sleep problems (e.g. asthma, chronic obstructive pulmonary disease, sleep apnea), parasomnias (e.g. sleepwalking) and sleep pattern disturbances, in keeping with the non-specific UK Biobank self-reported insomnia question. Duplicate codes were removed. This resulted in a final list of 181 Read codes (Table S1). Many Read terms for insomnia do not differentiate between diagnoses and symptoms, therefore we did not examine the prevalence of diagnoses alone^29^.

We mapped the BNF (British National Formulary) prescription codes in the TPP UK Biobank primary care data to the standard BNF codes produced by the NHS Business Services Authority (NHSBSA). The first six digits of the TPP BNF codes correspond to the first six digits of standard BNF codes (these relate to the BNF chapter, section and paragraph)^28^. We therefore reformatted the TPP BNF codes in our dataset to consist of six digit codes. We defined a prescription for insomnia symptoms as a prescription for a hypnotic medication (Six digit BNF code: 040101) (Table S2).

### 2.3 Covariates

To ascertain the characteristics of participants with insomnia symptoms identified by the self-report data and primary care records, we included a number of sociodemographic (age, sex, ethnic group, household income, Index of Multiple Deprivation, current employment status, highest qualification, household size, living with spouse/partner and home area population density), sleep (duration, chronotype, snoring, dozing, napping, how easy find getting up in the morning, night shift work), lifestyle (physical activity, tea/coffee intake, smoking, alcohol intake, risk taking) and health (BMI, menopause, depression, worrying, overall health rating) factors in our analysis. Further details on how the covariates were handled are provided in the Supplementary Method.

### 2.4 Statistical Analysis

We calculated the prevalence of insomnia symptoms in both the self-reported and primary care data. We also identified the proportion of self-reported insomnia symptom cases that were primary care insomnia symptom cases and vice versa. To identify and compare the characteristics of self-reported and primary care insomnia symptom cases, insomnia prevalence was stratified by sociodemographic, lifestyle, sleep and health variables, and visualised in coefficient plots. Analyses were performed in Stata version 16 via JupyterLab in DNA Nexus. Full code is available at https://github.com/MeldeLange/insomnia-comparison-study

### 2.5 Sensitivity Analyses

To explore the effect of using different definitions of insomnia symptoms in primary care on the number of cases and their overlap with self-reported insomnia symptom cases, we performed two sensitivity analyses. Firstly, we looked at the timing of symptoms, defining primary care cases as only those with a Read code in the 12 months or 4 weeks prior to UK Biobank baseline assessment. Secondly, we defined primary care cases as those with a hypnotic prescription prior to baseline or those with a Read code and concomitant prescription for hypnotics within 90 days of the Read code.

## 3 RESULTS

### 3.1 Participant Characteristics and Insomnia Symptom Prevalence

Characteristics of the study population, overall and stratified by self-reported or primary care insomnia symptoms case, are presented in Table 1 (see Supplemental Table S3 for full participant characteristics). In this study 45% of participants were male, 62% were aged 55 or over and 75% had a sleep duration of 7 hours or more. We found that 29% of the sample self-reported having insomnia symptoms, whilst only 6% had a primary care Read code for insomnia symptoms.

**TABLE 1.**
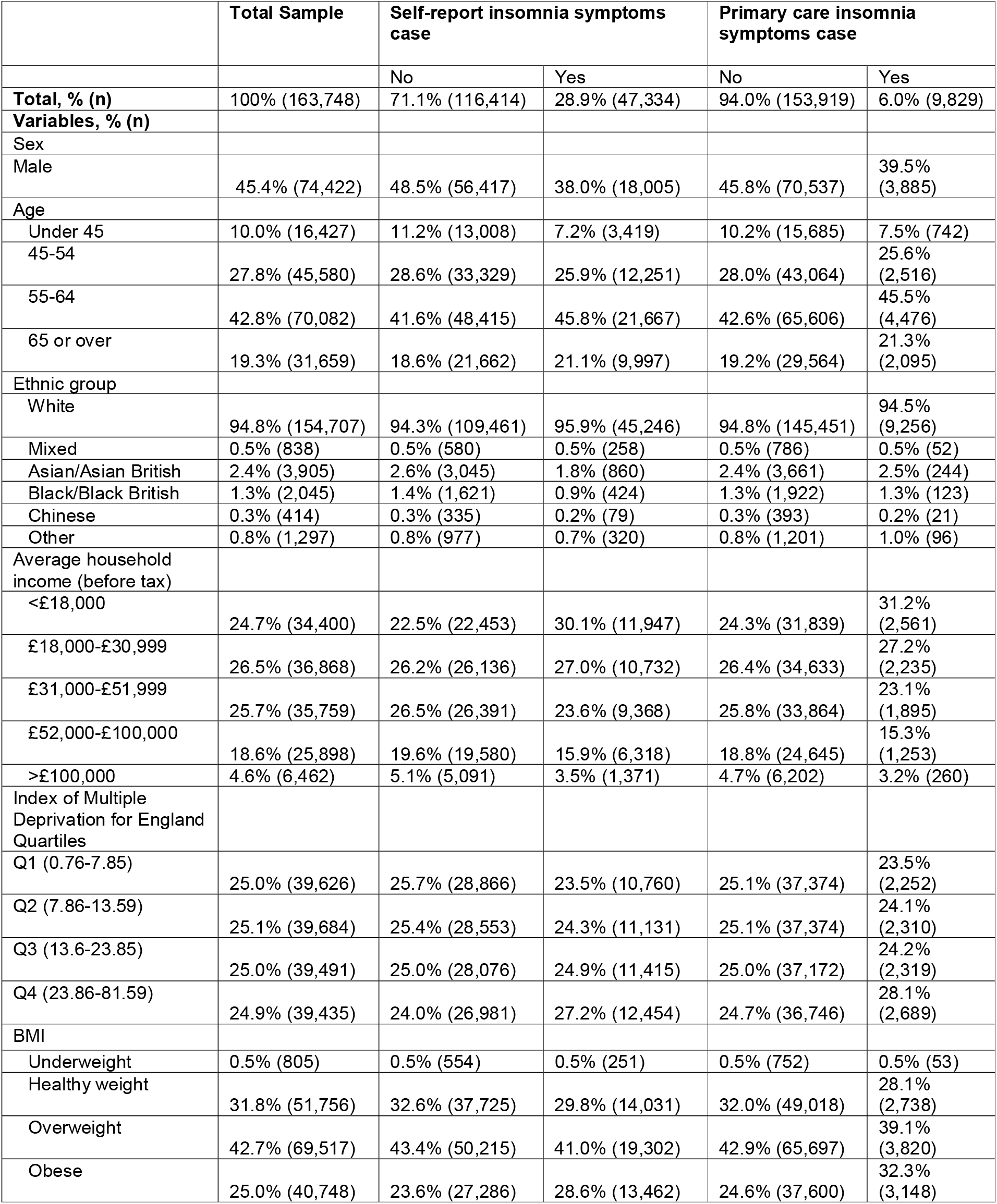
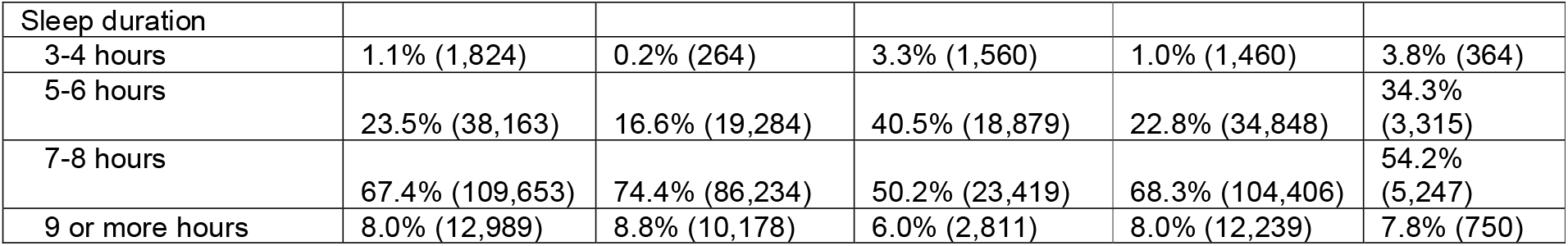
Characteristics of Total Sample and Groups Stratified by Insomnia Symptoms Status.

We found that only 10% of self-reported insomnia symptom cases were also a primary care insomnia symptom case. Meanwhile, only 49% of primary care insomnia symptom cases were also a self-reported case (Table 2).

**TABLE 2.**
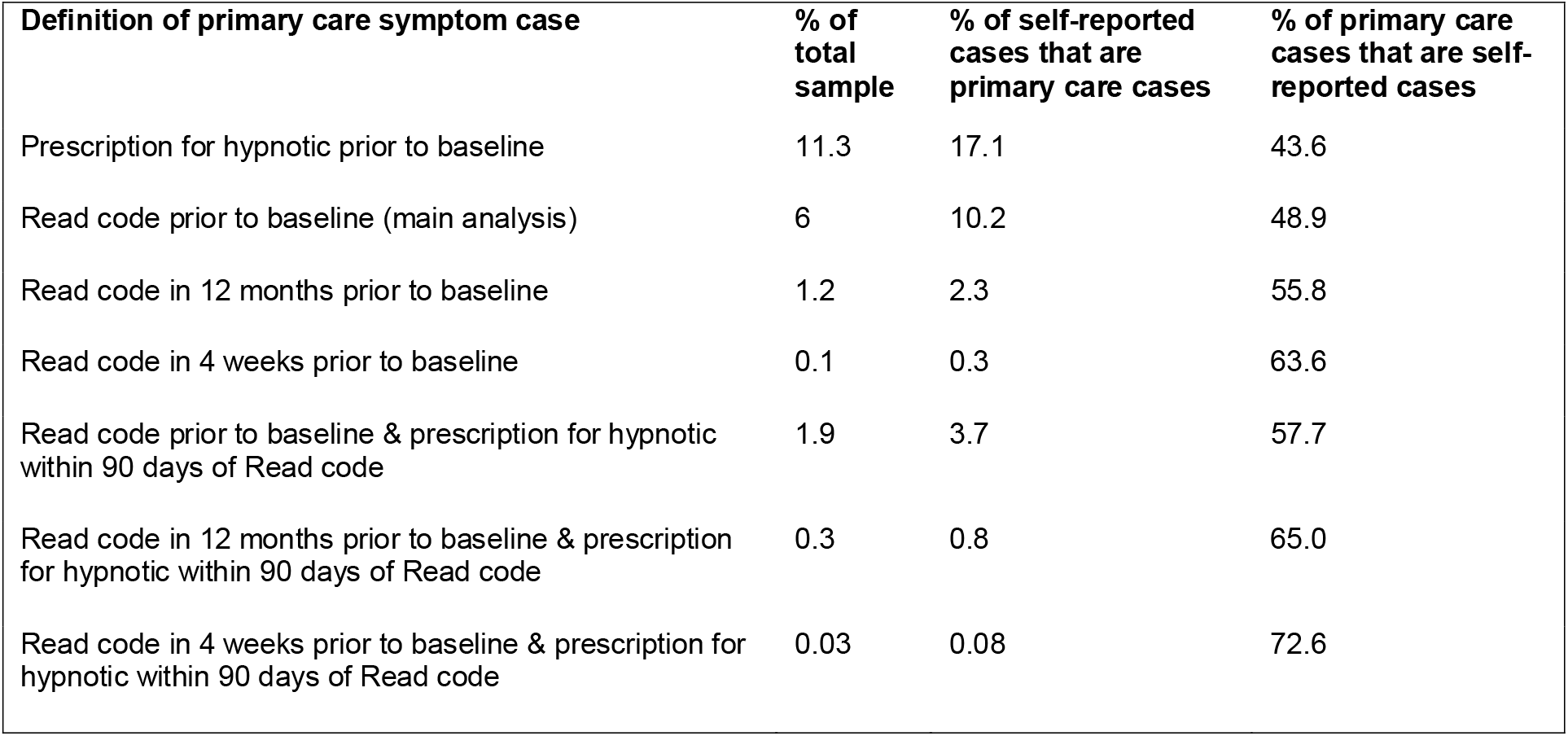
Cross-tabulation of Primary Care and Self-Reported Insomnia Symptom Cases.

### 3.2 Characteristics of self-reported and primary care insomnia symptom cases

Table S4 and Figures S1-4 show that participants who self-reported insomnia symptoms and those with insomnia Read codes in their primary care records had similar characteristics. Sociodemographic correlates of being a primary care or self-reported insomnia symptoms case included being female, older, in the lowest household income category (<£18,000), in the highest quartile of deprivation, in the “other” employment status category, with no qualifications, living in a one-person household, not living with a spouse/partner and not living in a rural area.

In addition, reporting a sleep duration of 3-4 hours, being a definite evening or definite morning chronotype, “often” dozing during the day, “usually” napping during the day, finding getting up in the morning “not at all easy” and not doing shift work were also characteristics of those reporting insomnia or having a primary care record indicating insomnia.

Lifestyle characteristics of primary care or self-reported insomnia symptoms cases included being in the lowest quartile of MET minutes/week, drinking 0-1 or 6+ cups of coffee per day, drinking 6+ cups of tea per day, being a previous or current smoker, and drinking alcohol less than once a month. Furthermore, health correlates of self-reported and primary care-recorded insomnia symptoms cases included women having been through the menopause, being underweight or obese, having experienced a depressed mood nearly every day in the past two weeks, being a worrier and rating your overall health as poor.

There were a few noticeable differences between the characteristics of self-reported and primary care insomnia symptom cases. Being a snorer was a correlate of being a primary care insomnia symptoms case, but being a non-snorer was a correlate of self-reporting insomnia. In addition, describing yourself as a risk taker was a characteristic of those having a primary care record indicating insomnia, whereas the opposite was true for self-reported cases. Differences in terms of ethnicity were also observed: being mixed or white was a correlate of being a self-reported case, whereas being in the ‘other’ category was a correlate of being a primary care case.

### 3.3 Sensitivity Analyses

Prevalence of primary care measured insomnia symptoms decreased from 6% to 0.1% of our sample when our definition of a primary care symptom case changed from having had an insomnia Read code prior to baseline to having had an insomnia Read code in the four weeks prior to baseline (Table 3). It also fell to 1.9% when our definition of a primary care case required having a Read code prior to baseline and being prescribed a hypnotic within 90 days of the Read code, and fell further to 0.03% when we required a Read code in the four weeks prior to baseline and being prescribed a hypnotic within 90 days of the Read code. The prevalence of being prescribed a hypnotic medication prior to baseline was higher than the prevalence of having an insomnia Read code prior to baseline (11% vs. 6%).

**TABLE 3.**
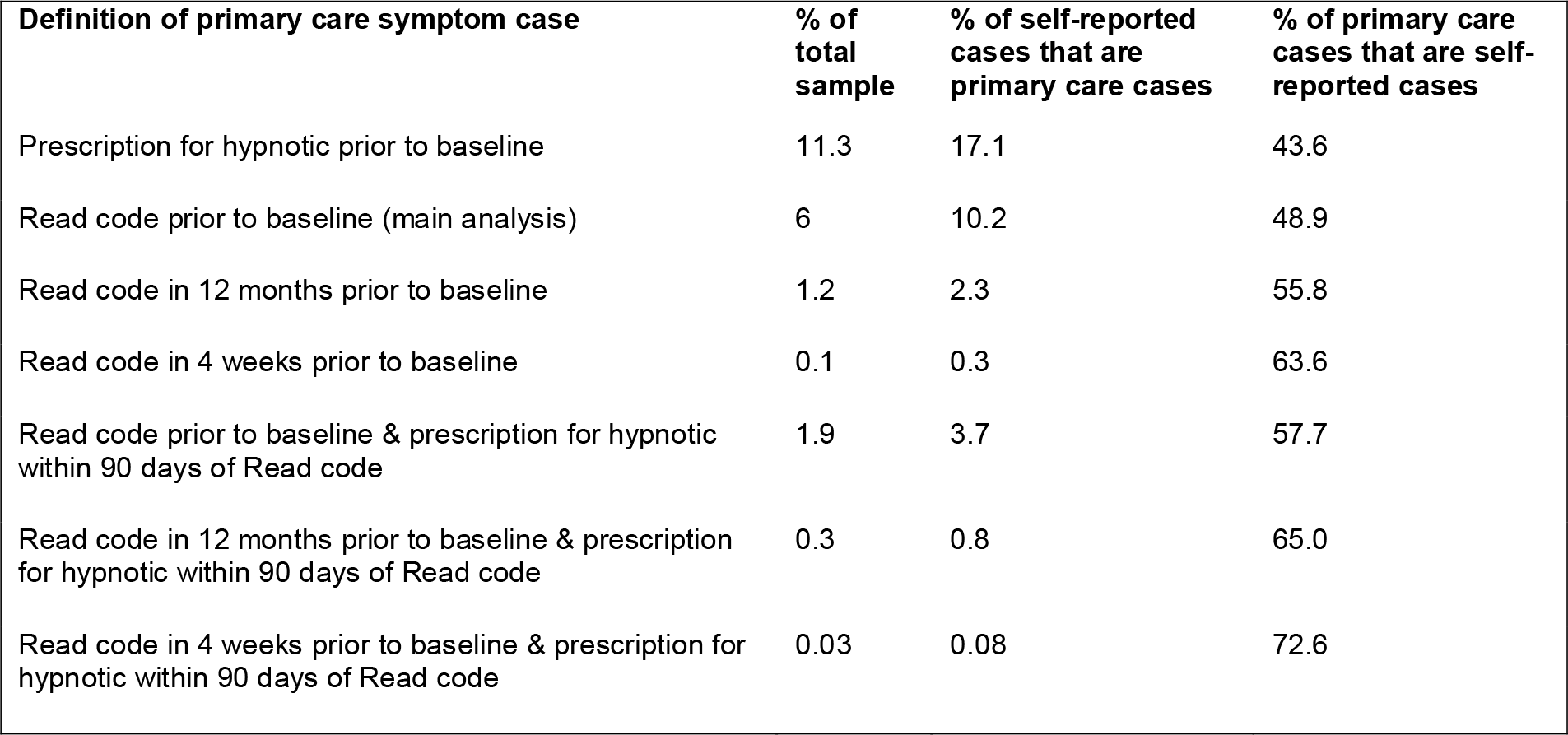
Prevalence of primary care measured insomnia symptoms cases and overlap with self-reported insomnia symptom cases according to different definitions of a primary care insomnia symptoms case.

As the strictness of our definition of a primary care insomnia symptom case increased, the proportion of primary care cases that were also self-reported cases (the specificity) increased and the proportion of self-reported cases that were also primary care cases decreased (the sensitivity) (See Tables S5-10 for full cross-tabulations).

## 4 DISCUSSION

We used data from the UK Biobank to compare the prevalence of self-reported insomnia symptoms to the prevalence indicated by linked primary care records.

Insomnia symptoms were common: 29% of our sample self-reported having frequent insomnia symptoms. This finding is highly consistent with a previous review of 50 studies from different countries, which estimated that around a third of the general population report having insomnia symptoms^15^. In this study only 6% of participants’ primary care records contained Read codes for insomnia symptoms. This is slightly lower than expected given that previous studies estimate that 6% of the general population meets the clinical criteria for an insomnia diagnosis^15^ and our study of symptoms was broader in scope. This inconsistency may be due to previous studies reporting insomnia diagnosis prevalence from interviews, whereas we measured the proportion of people who had acted on their symptoms and reported them to a primary care doctor. Our estimate is also slightly lower than a previous study by Klingman & Sprey (2020) which found the prevalence of insomnia diagnoses alone in primary care records in the US to be 9%^21^. This difference could be due to the difference in sample sizes (n=163,748 vs n=7,928), cultural differences in visiting a health practitioner for sleep-related issues, sample selection bias in either sample or true differences in the rates of insomnia in the two populations.

In our sample, 1.9% of participants had a Read code for insomnia symptoms and a prescription for a hypnotic medication within 90 days of that Read code. Klingman & Sprey (2020) reported a 4% prevalence of insomnia in the US with both diagnosis and prescription codes in primary care records. However, that study did not restrict when the medication was prescribed. Our finding that 11% of our sample had been prescribed a hypnotic medication was slightly higher than Klingman & Sprey’s prevalence of 7.6%. This could be due to cultural differences in prescribing practices and the availability of non-pharmaceutical treatments. The fact that we found prevalence of hypnotic prescriptions to be higher than the prevalence of insomnia symptom Read codes may be due to practitioners prescribing these drugs for conditions other than insomnia (such as agitation in dementia/psychotic disease or as a muscle relaxant for back pain) or prescribing them whilst documenting insomnia in the free text notes rather than recording a Read code.

We found that only 10% of people self-reporting insomnia symptoms had a primary care Read code for insomnia symptoms. This provides further evidence that only a small proportion of people experiencing insomnia seek help from a healthcare professional^22-26^, and suggests that EHRs only capture a small proportion of those experiencing insomnia symptoms. As these are likely to be the most severe cases, or those not responsive to self-management, this may lead to amplification bias in associations between insomnia and health outcomes such as cardiovascular disease.

The low level of help-seeking behaviour for insomnia may be because people perceive insomnia as something that is harmless, trivial, or amenable to self-management^30, 31^. In addition, stigma may deter people from seeking help^30-32^ or they may be unaware of the treatment options for insomnia, or concerned about the effectiveness and safety of sleeping tablets^30^. In England, although referral for cognitive behavioural therapy for insomnia (CBT-I) is recommended as a first line treatment when insomnia symptoms are unlikely to resolve soon, its availability is limited^33, 34^. Consequently, many people rely on self-help remedies such reading, listening to music and relaxation, or use over-the-counter or complementary and alternative medicine (CAM) therapies to aid sleep^23, 24, 31^. However, new digital treatments for insomnia, such as the NICE-approved Sleepio, could help to expand the treatment options available to primary care doctors^33^.

Surprisingly, we found that only 49% of primary care insomnia symptom cases also self-reported having insomnia symptoms. Possible explanations for this include that we looked at people’s primary care records from birth until they entered the UK Biobank study. It is possible that people may have experienced insomnia and visited their doctor a long time ago, then subsequently experienced an improvement in their symptoms before self-reporting their symptoms at the time of study entrance. This is supported by the fact that when we only included those with insomnia Read codes in the four weeks before baseline in our sensitivity analyses, the proportion of primary care insomnia symptom cases that self-reported insomnia symptoms rose (to 64% or 73% for those with a Read code 4 weeks prior to baseline and prescription within 90 days of the Read code). It is also possible that people with insomnia did not self-report having symptoms because they were ameliorated by medication or due to the stigma attached to having insomnia.

We found that the characteristics of self-reported and primary care-defined insomnia symptom cases were remarkably similar. Following previous studies^15, 21^, we found that key correlates of being a primary care or a self-reported insomnia symptom case were being female, older, not living with a partner, having lower educational attainment and incomes, and having poorer physical and mental health. We also found that women who had been through menopause, not living in a rural area, having an extreme chronotype, being less physically active, drank lots of tea or coffee, and smoking were predictors of primary care and self-reported insomnia symptom cases. These consistent findings suggest that primary care records can provide valuable evidence about population level risk factors for insomnia. They could also be clinically important for GPs as some of the characteristics identified (e.g. tea drinking / exercise) are modifiable lifestyle factors.

We also found that snoring predicted being a primary care insomnia symptoms case. However, the opposite was true of self-reported cases (i.e. self-reported insomniacs were less likely to report snoring). This suggests that snoring may be a key risk factor for prompting insomniacs to visit their primary care doctor to discuss their sleep. This may be because snoring can affect the partners’ sleep^35^ and mental health^36^, and consequently, their partner encourages them to seek medical help. However, this was not supported by our analysis which suggests that primary care insomnia cases are more likely to be living alone. More research is needed to understand the links between insomnia and other sleep related phenotypes.

Strengths of our study include the large sample size, which meant our estimates, even within strata, were very precise. In addition, the extensive, detailed self-report questionnaire combined with linked EHR data allowed us to explore, validate and triangulate across multiple definitions of insomnia. Our study also has several limitations. Firstly, the UK Biobank is not representative of the UK population, with participants more likely to be female, healthier, older and live in less socioeconomically deprived areas than non-participants^27^. If having insomnia also affects participation in the UK Biobank, then this may have caused selection bias in our estimates of insomnia prevalence.

A further limitation is the lack of a Gold Standard measure of insomnia. In this study, insomnia prevalence differed depending on the primary care definition used and our estimates differed from those of previous research, which again may be due to differences in definition. This makes it difficult to compare the prevalence of insomnia across populations. The prevalence of insomnia cases was extremely low when we placed time restrictions on insomnia Read codes or when a concomitant hypnotic prescription was required. This suggests that the most constructive measure of insomnia in primary care data is having a Read code for insomnia symptoms alone, at any point throughout a person’s medical history. It should also be noted that the self-report insomnia question asked in the UK Biobank did not encompass early morning awakenings or impaired daytime function. Our definition of self-reported insomnia symptoms is therefore not in line with established guidelines for insomnia diagnosis and treatment^37^.

In conclusion, this study found that in a large sample, primary care symptom codes only capture a small proportion of those experiencing insomnia symptoms in the population. As these are likely to be the most extreme cases, associations between insomnia and other health outcomes may be amplified in primary care data. Nonetheless, EHRs provide a valuable data source for studying insomnia, offering objective insights into severe insomnia, large sample sizes and longitudinal data. Furthermore, the relationships observed between insomnia symptoms and socio-demographic characteristics were consistent in both self-report and primary care datasets. Consequently, researchers exploring population-level risk factors for insomnia are likely to draw similar conclusions using either dataset. Further studies should replicate our findings in other populations and examine the best ways to treat insomnia in primary care.

## Supporting information

Figures S1-4 Coefficient Plots

Table S1 Read Codelist

Table S2 Hypnotic Drugs

Table S3 Sample Characteristics

Table S4 Stratification Table

Tables S5-10 Sensitivity Analyses

## Data Availability

The UK Biobank dataset used to conduct the research in this paper is available via application directly to the UK Biobank. Applications are assessed for meeting the required criteria for access, including legal and ethics standards. More information regarding data access can be found at https://www.ukbiobank.ac.uk/enable-your-research. Full code for all analyses is available at https://github.com/MeldeLange/insomnia-comparison-study

## Acknowledgements

This research has been conducted using the UK Biobank Resource under Application Number 88626.

