## Supplementary material for "Insomnia symptom prevalence in England: A comparison of self-reported data and primary care records in the UK Biobank": Figures S1-4 Coefficient Plots

**FIGURE S1** Coefficient plot of primary care and self-reported insomnia symptom cases stratified by sex, age, ethnicity, household income, deprivation and employment status

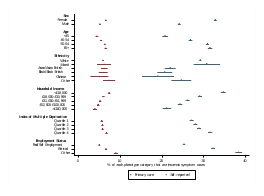

**FIGURE S2** Coefficient plot of primary care and self-reported insomnia symptom cases stratified by qualifications, household size, living with spouse/partner, population density, sleep duration, chronotype and snoring

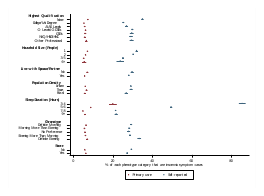

**FIGURE S3** Coefficient plot of primary care and self-reported insomnia symptom cases stratified by daytime dozing, daytime napping, ease of getting up in the morning, working night shifts, MET mins/week, and coffee and tea intake.

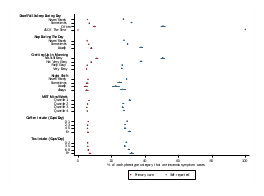

**FIGURE S4** Coefficient plot of primary care and self-reported insomnia symptom cases stratified by BMI, risk taking, smoking status, alcohol intake, menopause, depression, worrier and overall health rating

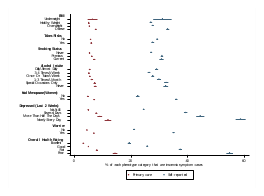
