## Supplementary Methods for "Insomnia symptom prevalence in England: A comparison of self-reported data and primary care records in the UK Biobank"

**SUPPLEMENTARY METHOD**

**Handling of covariates**

Data on most covariates was self-reported by participants through a touchscreen questionnaire completed at their baseline UK Biobank assessment centre visit (Questionnaire available at https:/ /biobank.ctsu.ox.ac.uk/showcase/ukb/docs/TouchscreenQuestionsMainFinal.pdf). Body mass index (kg/m^2^) was calculated from height and weight measurements taken during the same visit.

We collapsed continuous and discrete variables to make ordinal variables in order to facilitate stratifying insomnia symptom prevalence by our covariates.

**Sociodemographic factors**

Sex (male/female) was taken from the central NHS registry at recruitment, but in some cases was updated by participants.

We collapsed ethnic group from 18 to six categories: White, Mixed, Asian/Asian British, Black/Black British, Chinese and Other.

Age when attended assessment centre was derived from date of birth and date of attending baseline assessment centre, then truncated to whole year part. We collapsed this to four age categories: Under 45, 45-54, 55-64 and 65+.

Average household income (before tax) consisted of five categories: Less than £18,000/£18,000-£30,999/£31,000-£51,999/£52,000-£100,000/Greater than £100,000. We coded “Do not know” and “Prefer not to answer” as missing.

The Index of Multiple Deprivation for England (English Indices of Deprivation 2010) is a continuous variable measuring multiple deprivation at the small area level. We split this into quartiles to create an ordinal variable with four categories.

We simplified current employment status from 9 to 3 categories. Participants could select more than one answer. Anyone that selected “employed” or “self-employed” in any answer was recoded as “Employed”, anyone who selected “Retired” in any answer was recoded as “Retired”. Participants not falling in the above two categories (including those selecting "None of the above") were included in a third ‘Other’ category. Those who selected “prefer not to answer” were coded as missing.

We derived our highest qualification variable from participants being asked which of a set of qualifications they have. They could select more than one answer. We recoded participants’ answers to reflect the highest qualification a person had. For example, anyone who selected “College/University degree” at all was coded as their highest qualification being a “College/University degree”. If they did not have a degree but did select “A/AS levels of equivalent” they were coded as their highest qualification being “A/AS levels”. Those selecting “None of the above” were coded as having “None”. Those selecting “Prefer not to answer” were recoded as missing. This resulted in an ordinal highest qualifications variable with 7 categories: “College or university degree”, “A levels, AS levels or equivalent”, “O levels, GCSEs or equivalent”, “CSEs or equivalent”, “NVQ or HND or HNC or equivalent”, “Other professional qualifications”, “None”.

Our “household size” variable was derived from participants’ answers to the question: “"Including yourself, how many people are living together in your household? (Include those who usually live in the house such as students living away from home during term, partners in the armed forces or professions such as pilots)". We recoded those selecting “Prefer not to answer” or “Do not know” as missing. We then collapsed the discrete variable into four categories: “1 person”, “2 people”, “3-5 people” and “6+ people”.

Our “Live with spouse/partner” variable was derived from participants’ answers to how people in the household were related to the participant (9 categories), which were then collapsed into a binary variable where living with “husband, wife or partner” was coded as “Yes” and all other categories were coded as “No”.

Home area population density - urban or rural (17 categories) was collapsed into a categorical variable with three categories. All urban categories were coded as “Urban”, all categories mentioning a town were coded as “Town” and all categories mentioning villages, hamlets, isolated dwellings or rural areas were coded as “Rural”. Those coded as “Postcode not linkable” were recoded as missing. We found we had a very small number of participants living in areas of Scotland according to this variable. These people have English primary care records. Therefore they must have lived in England at some point but had moved to Scotland by the time of the initial UK Biobank assessment centre visit, or could potentially live close to the English border so are registered with an English GP. We coded these participants as having missing data just for this variable.

**Sleep factors**

We derived our sleep duration variable from answers to the question "About how many hours sleep do you get in every 24 hours? (please include naps)". We recoded those selecting “Prefer not to answer” or “Do not know” as missing. We recoded extreme values of sleep duration (over 18 hours and under three hours) to missing. We then collapsed the discrete variable into four categories “3-4 hours”, “5-6 hours”, “7-8 hours” and “9 or more hours”.

We derived our chronotype variable from participants being asked whether they considered themselves to be: “Definitely a morning person”, “More a morning than evening person”, “More an evening than a morning person”, “Definitely an evening person”, “Do not know” or “Prefer not to answer”. We coded “Prefer not to answer” as missing and “Do not know” as “No preference”. From this we derived a five point ordinal variable for chronotype: “Definite morning”, “More a morning than evening”, “No Preference”, “Evening more than morning” and “Definite evening”.

We derived our snoring variable from participants being asked "Does your partner or a close relative or friend complain about your snoring?". We coded “Prefer not to answer” and “Do not know” as missing to create a binary (Yes/No) variable.

We derived our dozing/sleeping during the day variable from participants being asked "How likely are you to doze off or fall asleep during the daytime when you don't mean to? (e.g. when working, reading or driving)". We coded “Prefer not to answer” and “Do not know” as missing to create an ordinal variable with four categories: "Never/rarely", "Sometimes", "Often" and "All of the time".

Participants were asked whether they nap during the day. We coded “Prefer not to answer” as missing to create a 3 category ordinal variable: "Never/rarely", "Sometimes" and “Usually”.

We derived our how easy find getting up in the morning variable from participants being asked "On an average day, how easy do you find getting up in the morning?". We coded “Prefer not to answer” and “Do not know” as missing to create an ordinal variable with four categories: "Not at all easy", "Not very easy", "Fairly easy" and "Very easy".

Participants who stated in a previous question that their job involves shift work were then asked whether their job involves night shift work. To derive our night shift variable we coded “Prefer not to answer” and “Do not know” as missing. We recoded anyone with missing data (i.e. who wasn’t asked the question because they said they didn’t do shift work) as “Never/rarely”. This gave us a four category ordinal variable: “Never/rarely", "Sometimes", "Usually" and "Always".

**Lifestyle factors**

Our Metabolic Equivalent Task (MET) minutes per week variable was derived from the continuous variable Total Metabolic Equivalent Task (MET) minutes per week for all activity including walking, moderate and vigorous activity. We split this into quartiles to create an ordinal variable with four categories.

Participants were asked "How many cups of coffee do you drink each DAY? (Include decaffeinated coffee)". To derive our coffee intake variable we coded “Prefer not to answer” and “Do not know” as missing. We coded “Less than one cup” as zero cups. We then collapsed the discrete variable into four categories: “0-1 cups per day”, “2-3 cups per day”, “4-5 cups per day” and “6 or more cups per day”.

Participants were asked "How many cups of tea do you drink each DAY? (Include black and green tea)". To derive our coffee intake variable we coded “Prefer not to answer” and “Do not know” as missing. We coded “Less than one cup” as zero cups. We then collapsed the discrete variable into four categories: “0-2 cups per day”, “3-5 cups per day”, “6-8 cups per day” and “9 or more cups per day”.

For our smoking status variable we coded “Prefer not to answer” as missing to create a three category ordinal variable: “Current”, “Previous”, “Never”.

To derive our alcohol intake frequency variable we coded “Prefer not to answer” as missing to create a six category ordinal variable: “Daily or almost daily”, “Three or four times a week”, “Once or twice a week”, “One to three times a month”, “Special occasions only” and “Never”.

Participants were asked "Would you describe yourself as someone who takes risks?". We coded “Prefer not to answer” and “Do not know” as missing to create a binary (Yes/No) variable.

**Health factors**

A continuous BMI (kg/m^2^) variable was constructed by UK Biobank from height and weight measured during the initial Assessment Centre visit. The value is missing if either height or weight readings were omitted. We recoded BMI into four categories based on WHO BMI categories: “Underweight” (<18.5), “Healthy weight” (18.5-24.9),”Overweight” (25-29.9) and “Obese” (over 30).

Females only were asked "Have you had your menopause (periods stopped)?". We coded “Not sure – had a hysterectomy” as “Yes”. We coded “Not sure – other reason” and “Prefer not to answer” as missing. This created a binary (Yes/No) menopause variable.

Participants were asked "Over the past two weeks, how often have you felt down, depressed or hopeless?" We coded “Prefer not to answer” and “Do not know” as missing to leave us with a four category ordinal variable: “Not at all", "Several days", "More than half the days" and "Nearly every day".

Participants were asked "Are you a worrier?". We coded “Prefer not to answer” and “Do not know” as missing to create a binary (Yes/No) worrier variable.

Participants were asked "In general how would you rate your overall health?" We coded “Prefer not to answer” and “Do not know” as missing to give us a four category ordinal variable: “"Excellent", "Good", "Fair" and "Poor".
