## Supplementary material for "Insomnia symptom prevalence in England: A comparison of self-reported data and primary care records in the UK Biobank": Table S1 Read Codelist

**Table S1: Insomnia Symptoms Read Code List**

| **Read Code** | **Description** |
| --- | --- |
| 1B1B. | insomnia (& c/o) |
| 1B1B0 | initial insomnia |
| 1B1B1 | middle insomnia |
| 1B1B2 | late insomnia |
| 3148. | sleep studies - procedure |
| 663N. | asthma disturbing sleep |
| 663N1 | asthma disturbs sleep weekly |
| 663N2 | asthma disturbs sleep frequently |
| E274. | non-organic sleep disorders (& [hypersomnia] or [insomnia]) |
| E2740 | unspecified non-organic sleep disorder |
| E2741 | insomnia: [transient] or [nos] |
| E2742 | persistent insomnia |
| E2747 | sleepwalking |
| E2748 | sleep terrors |
| E274A | sleep drunkenness |
| E274B | repeated rapid eye movement sleep interruptions |
| E274C | other sleep stage or arousal dysfunction |
| E274D | (repetitive intrusions of sleep) or (restless sleep) |
| E274E | "short-sleeper" |
| E274F | reversed sleep-wake cycle |
| E274y | (dreams) or (other non-organic sleep disorder) |
| E274z | non-organic sleep disorder nos |
| Eu510 | nonorganic insomnia |
| Eu512 | [x] nonorganic disorder of the sleep-wake schedule: [psychogenic inversion of circadian rhythm (including nyctohemeral rhythm and sleep rhythm)] |
| Eu51y | [x]other nonorganic sleep disorders |
| Eu51z | [x]nonorganic sleep disorder, unspecified |
| Fy00. | disorders of initiating and maintaining sleep |
| Fy03. | sleep apnoea (& [obstructive]) |
| Fyu58 | [x]other sleep disorders |
| K5A21 | menopausal sleeplessness |
| R005. | [d]sleep disturbances |
| R0050 | [d]sleep disturbance, unspecified |
| R0051 | [d]insomnia with sleep apnoea |
| R0052 | [d]insomnia nos |
| R0053 | ([d]hypersomnia with sleep apnoea) or (sleep apnoea syndrome) |
| R0055 | [d]sleep rhythm inversion |
| R0056 | [d]sleep rhythm irregular |
| R0057 | [d]sleep-wake rhythm non-24-hour cycle |
| R0058 | [d]sleep dysfunction with sleep stage disturbance |
| R0059 | [d]sleep dysfunction with arousal disturbance |
| R005z | [d]sleep dysfunction nos |
| TJ7z0 | adverse reaction to sleeping pill nos |
| TK02. | [x]overdose - sleeping tabs |
| U1A2. | [x]accidental poisoning with sleeping tablets |
| U202. | [x]overdose - sleeping tabs |
| Ua15W | ability to sleep |
| Ua1FM | sleep and rest interventions |
| Ua1ZQ | sleep talking |
| X003N | familial fatal insomnia |
| X007q | sleep-wake disorder |
| X007s | insomnia nos |
| X007u | difficulty getting to sleep |
| X007v | difficulty in sleep maintenance |
| X0083 | sleep apnoea |
| X0084 | obstructive sleep apnoea |
| X0085 | central sleep apnoea |
| X0086 | mixed sleep apnoea |
| X0087 | alveolar sleep apnoea |
| X0088 | transient sleep-wake rhythm disorder |
| X0089 | delayed sleep phase syndrome |
| X008A | non-24 hour sleep-wake cycle |
| X008C | sleep-related head banging |
| X008D | sleep-related painful erections |
| X764D | low level of awareness whilst sleep walking |
| X764E | low level of reactivity whilst sleep walking |
| X764F | low level of motor skill whilst sleep walking |
| X764G | blank, staring face whilst sleep walking |
| X764H | unresponsive to communication whilst sleep walking |
| X764I | not easily wakened from sleep walking |
| X764J | no recollection of sleep walk |
| X764K | sleep automatism |
| X769S | wakefulness |
| X76AE | sleep rhythm problem |
| X76AF | cannot sleep at all |
| X76AG | not getting enough sleep |
| X76AJ | wakes and cannot sleep again |
| X76AK | wakes early |
| X76AL | circumstances interfere with sleep |
| X76AM | symptoms interfere with sleep |
| X76AN | restless sleep |
| X76AO | unrefreshed by sleep |
| X77f4 | sleep latency test |
| X77f5 | full sleep study |
| X77f6 | mini-sleep study |
| XE0ux | insomnia (& symptom) or somnolence |
| XE1Yg | transient insomnia |
| XE1Yi | repetitive intrusions of sleep |
| XE1Yj | other non-organic sleep disorder |
| XE1Zr | non-organic disorder of the sleep-wake schedule |
| XE1bI | sleep disorders (& [insomnia] or [nightmares] or [sleepwalking (& somnambulism)]) |
| XE1gP | [d]sleep dysfunction nec |
| XE2Pv | insomnia |
| XE2Q5 | non-organic sleep disorder |
| XE2cd | [d]sleep disturbances (& [hypersomnia] or [insomnia]) |
| XE2nU | [d]hypersomnia with sleep apnoea |
| XM06i | sleep disturbance |
| XM06j | irregular sleep-wake pattern |
| XM06k | disorder of sleep-wake cycle |
| XM0CT | c/o - insomnia |
| XM0Go | appliance for sleep apnoea |
| XM0yu | [d]insomnia |
| XSGLz | light sleep |
| Xa7wV | difficulty sleeping |
| XaEGP | [d]sleep apnoea syndrome |
| XaFqr | poor sleep pattern |
| XaIp8 | sleep management |
| XaIti | delayed onset of sleep |
| XaJKk | sleep studies - specialty |
| XaKv8 | chronic obstructive pulmonary disease disturbs sleep |
| XaMs7 | sleep studies nec |
| XaN5q | seen in sleep clinic |
| XaO8S | pittsburgh sleep quality index |
| XaO8T | insomnia severity index |
| XaOb7 | referral to sleep clinic |
| XaP4v | sleep hygiene behaviour education |
| XaP4w | sleep restriction therapy |
| XaYGN | able to sleep with sedation |
| Xaag2 | on melatonin for sleep disorder |
| XabE4 | symptom assessment scale - difficulty sleeping score |
| XafMO | signposting to the sleep council |
| ZV1B1 | [v]personal history of unhealthy sleep-wake schedule |
| c88A. | sleepia 50mg capsule |
| c88G. | vantage pharmacy sleep aid 50mg tablet |
| c88H. | care night time sleep aid 25mg tablet |
| iz1E. | natrasleep tablet |
| x02kT | natrasleep |
| x03mt | sleepia |
| x05t9 | vantage pharmacy sleep aid |
| .1B1B | insomnia (& symptom) or somnolence |
| .1B1Q | poor sleep pattern |
| .1BX0 | delayed onset of sleep |
| .1BX9 | light sleep |
| .38D0 | pittsburgh sleep quality index |
| .38D1 | insomnia severity index |
| .663N | asthma disturbing sleep |
| .66Yg | chronic obstructive pulmonary disease disturbs sleep |
| .8G99 | sleep restriction therapy |
| .8G9B | sleep hygiene behaviour education |
| .8HTn | referral to sleep clinic |
| .8Q0. | sleep management |
| .9Nk0 | seen in sleep clinic |
| .E4A. | sleep disorders (& [insomnia] or [nightmares] or [sleepwalking (& somnambulism)]) |
| .E4A0 | sleep terrors |
| .E4A1 | wakes early |
| .H66. | sleep apnoea |
| .R05. | [d]sleep disturbances (& [hypersomnia] or [insomnia]) |
| .R050 | [d]sleep disturbance, unspecified |
| .R051 | [d]insomnia with sleep apnoea |
| .R052 | [d]insomnia |
| .R053 | [d]hypersomnia with sleep apnoea |
| .R055 | [d]sleep rhythm inversion |
| .R056 | [d]sleep rhythm irregular |
| .R057 | [d]sleep-wake rhythm non-24-hour cycle |
| .R058 | [d]sleep dysfunction with sleep stage disturbance |
| .R059 | [d]sleep dysfunction with arousal disturbance |
| .R05Z | [d]sleep dysfunction nec |
| 1B1Q. | poor sleep pattern |
| 1BX0. | delayed onset of sleep |
| 38D0. | pittsburgh sleep quality index |
| 38D1. | insomnia severity index |
| 38Da. | berlin questionnaire for sleep apnoea |
| 66Yg. | chronic obstructive pulmonary disease disturbs sleep |
| 70658 | sleep studies - procedure |
| 7065A | sleep studies nec |
| 745A2 | mini-sleep study |
| 7P1B0 | full sleep study |
| 8G99. | sleep restriction therapy |
| 8G9B. | sleep hygiene behaviour education |
| 8HTn. | referral to sleep clinic |
| 8Q0.. | sleep management |
| 9Ngt. | on melatonin for sleep disorder |
| 9Nk0. | seen in sleep clinic |
| 9b9Y. | sleep studies - specialty |
| Eu51. | non-organic sleep disorder |
| Eu513 | sleepwalking |
| Eu514 | sleep terrors |
| Fy0.. | sleep-wake disorder |
| Fy02. | disorder of sleep-wake cycle |
| H5B.. | sleep apnoea |
| H5B0. | obstructive sleep apnoea |
| X007t | nonorganic insomnia |
