## Supplementary material for "Insomnia symptom prevalence in England: A comparison of self-reported data and primary care records in the UK Biobank": Table S2 Hypnotic Drugs

**Table S2: List of hypnotic drugs**

**Hypnotics included under six digit BNF code 040101**

Melatonin

Toquilone

Chloral hydrate

Clomethiazole

Flunitrazepam

Flurazepam

Loprazolam

Lormetazepam

Nitrazepam

Temazepam

Zaleplon

Triclofos sodium

Zolpidem

Zopiclone
