## Supplementary material for "Insomnia symptom prevalence in England: A comparison of self-reported data and primary care records in the UK Biobank": Table S3 Sample Characteristics

TABLE S3 Characteristics of Total Sample and Groups Stratified by Insomnia Symptoms Status

|  | **Total Sample** | **Self-report insomnia symptoms case** | | **Primary care insomnia symptoms case** | |
| --- | --- | --- | --- | --- | --- |
|  |  | No | Yes | No | Yes |
| **Total, % (n)** | 100% (163,748) | 71.1% (116,414) | 28.9% (47,334) | 94.0% (153,919) | 6.0% (9,829) |
| **Variables, % (n)** |  |  |  |  |  |
| Male | 45.4% (74,422) | 48.5% (56,417) | 38.0% (18,005) | 45.8% (70,537) | 39.5% (3,885) |
| Age |  |  |  |  |  |
| Under 45 | 10.0% (16,427) | 11.2% (13,008) | 7.2% (3,419) | 10.2% (15,685) | 7.5% (742) |
| 45-54 | 27.8% (45,580) | 28.6% (33,329) | 25.9% (12,251) | 28.0% (43,064) | 25.6% (2,516) |
| 55-64 | 42.8% (70,082) | 41.6% (48,415) | 45.8% (21,667) | 42.6% (65,606) | 45.5% (4,476) |
| 65 or over | 19.3% (31,659) | 18.6% (21,662) | 21.1% (9,997) | 19.2% (29,564) | 21.3% (2,095) |
| Ethnic group |  |  |  |  |  |
| White | 94.8% (154,707) | 94.3% (109,461) | 95.9% (45,246) | 94.8% (145,451) | 94.5% (9,256) |
| Mixed | 0.5% (838) | 0.5% (580) | 0.5% (258) | 0.5% (786) | 0.5% (52) |
| Asian/Asian British | 2.4% (3,905) | 2.6% (3,045) | 1.8% (860) | 2.4% (3,661) | 2.5% (244) |
| Black/Black British | 1.3% (2,045) | 1.4% (1,621) | 0.9% (424) | 1.3% (1,922) | 1.3% (123) |
| Chinese | 0.3% (414) | 0.3% (335) | 0.2% (79) | 0.3% (393) | 0.2% (21) |
| Other | 0.8% (1,297) | 0.8% (977) | 0.7% (320) | 0.8% (1,201) | 1.0% (96) |
| Average household income (before tax) |  |  |  |  |  |
| <£18,000 | 24.7% (34,400) | 22.5% (22,453) | 30.1% (11,947) | 24.3% (31,839) | 31.2% (2,561) |
| £18,000-£30,999 | 26.5% (36,868) | 26.2% (26,136) | 27.0% (10,732) | 26.4% (34,633) | 27.2% (2,235) |
| £31,000-£51,999 | 25.7% (35,759) | 26.5% (26,391) | 23.6% (9,368) | 25.8% (33,864) | 23.1% (1,895) |
| £52,000-£100,000 | 18.6% (25,898) | 19.6% (19,580) | 15.9% (6,318) | 18.8% (24,645) | 15.3% (1,253) |
| >£100,000 | 4.6% (6,462) | 5.1% (5,091) | 3.5% (1,371) | 4.7% (6,202) | 3.2% (260) |
| Index of Multiple Deprivation for England Quartiles |  |  |  |  |  |
| Q1 (0.76-7.85) | 25.0% (39,626) | 25.7% (28,866) | 23.5% (10,760) | 25.1% (37,374) | 23.5% (2,252) |
| Q2 (7.86-13.59) | 25.1% (39,684) | 25.4% (28,553) | 24.3% (11,131) | 25.1% (37,374) | 24.1% (2,310) |
| Q3 (13.6-23.85) | 25.0% (39,491) | 25.0% (28,076) | 24.9% (11,415) | 25.0% (37,172) | 24.2% (2,319) |
| Q4 (23.86-81.59) | 24.9% (39,435) | 24.0% (26,981) | 27.2% (12,454) | 24.7% (36,746) | 28.1% (2,689) |
| Current employment status |  |  |  |  |  |
| Paid employment / self-employed | 56.3% (91,752) | 59.1% (68,514) | 49.3% (23,238) | 56.8% (87,095) | 47.6% (4,657) |
| Retired | 34.7% (56,594) | 33.1% (38,330) | 38.7% (18,264) | 34.4% (52,806) | 38.7% (3,788) |
| Other | 9.0% (14,718) | 7.8% (9,067) | 12.0% (5,651) | 8.7% (13,386) | 13.6% (1,332) |
| Highest qualification |  |  |  |  |  |
| None | 18.2% (29,457) | 16.7% (19,173) | 22.0% (10,284) | 18.0% (27,321) | 22.0% (2,136) |
| College/University degree | 31.0% (50,109) | 32.6% (37,476) | 27.0% (12,633) | 31.2% (47,475) | 27.2% (2,634) |
| A/AS levels or equivalent | 10.7% (17,327) | 11.0% (12,675) | 9.9% (4,652) | 10.7% (16,350) | 10.1% (977) |
| O levels/GCSEs or equivalent | 21.9% (35,390) | 21.6% (24,846) | 22.5% (10,544) | 21.8% (33,226) | 22.3% (2,164) |
| CSEs or equivalent | 5.9% (9,502) | 5.8% (6,697) | 6.0% (2,805) | 5.9% (8,941) | 5.8% (561) |
| NVQ/HND/HNC or equivalent | 7.1% (11,436) | 7.0% (8,106) | 7.1% (3,330) | 7.1% (10,777) | 6.8% (659) |
| Other professional qualifications | 5.3% (8,613) | 5.3% (6,078) | 5.4% (2,535) | 5.3% (8,056) | 5.7% (557) |
| Household size |  |  |  |  |  |
| 1 person | 18.7% (30,323) | 17.8% (20,620) | 20.7% (9,703) | 18.4% (28,197) | 21.8% (2,126) |
| 2 people | 48.7% (79,181) | 47.7% (55,214) | 51.0% (23,967) | 48.6% (74,320) | 49.9% (4,861) |
| 3-5 people | 31.4% (51,012) | 33.1% (38,217) | 27.2% (12,795) | 31.6% (48,377) | 27.1% (2,635) |
| 6 or more people | 1.3% (2,071) | 1.4% (1,581) | 1.0% (490) | 1.3% (1,959) | 1.2% (112) |
| Live with spouse/partner | 90.1% (119,229) | 90.3% (85,848) | 89.6% (33,381) | 90.2% (112,525) | 88.0% (6,704) |
| Home area population density |  |  |  |  |  |
| Urban | 83.3% (134,995) | 83.1% (95,729) | 83.8% (39,266) | 83.4% (126,909) | 82.8% (8,086) |
| Town | 9.5% (15,424) | 9.5% (10,930) | 9.6% (4,494) | 9.5% (14,411) | 10.4% (1,013) |
| Rural | 7.2% (11,585) | 7.4% (8,490) | 6.6% (3,095) | 7.2% (10,923) | 6.8% (662) |
| Primary care insomnia case | 6.0% (9,829) | 4.3% (5,022) | 10.2% (4,807) | 0.0% (0) | 100.0% (9,829) |
| Self-report insomnia case | 28.9% (47,334) | 0.0% (0) | 100.0% (47,334) | 27.6% (42,527) | 48.9% (4,807) |
| Sleep duration |  |  |  |  |  |
| 3-4 hours | 1.1% (1,824) | 0.2% (264) | 3.3% (1,560) | 1.0% (1,460) | 3.8% (364) |
| 5-6 hours | 23.5% (38,163) | 16.6% (19,284) | 40.5% (18,879) | 22.8% (34,848) | 34.3% (3,315) |
| 7-8 hours | 67.4% (109,653) | 74.4% (86,234) | 50.2% (23,419) | 68.3% (104,406) | 54.2% (5,247) |
| 9 or more hours | 8.0% (12,989) | 8.8% (10,178) | 6.0% (2,811) | 8.0% (12,239) | 7.8% (750) |
| Chronotype |  |  |  |  |  |
| Definite morning | 24.5% (40,046) | 24.4% (28,348) | 24.8% (11,698) | 24.5% (37,509) | 25.9% (2,537) |
| Morning more than evening | 31.9% (51,985) | 32.2% (37,322) | 31.1% (14,663) | 32.0% (49,102) | 29.4% (2,883) |
| No preference | 10.6% (17,334) | 10.8% (12,494) | 10.3% (4,840) | 10.6% (16,329) | 10.3% (1,005) |
| Evening more than morning | 25.3% (41,229) | 25.4% (29,433) | 25.0% (11,796) | 25.2% (38,724) | 25.6% (2,505) |
| Definite evening | 7.7% (12,590) | 7.2% (8,394) | 8.9% (4,196) | 7.6% (11,719) | 8.9% (871) |
| Snore | 37.6% (57,300) | 38.4% (41,725) | 35.9% (15,575) | 37.5% (53,720) | 39.7% (3,580) |
| Doze/fall asleep during the day when don’t mean to |  |  |  |  |  |
| Never/rarely | 75.8% (123,494) | 77.6% (89,897) | 71.4% (33,597) | 76.1% (116,544) | 71.3% (6,950) |
| Sometimes | 21.3% (34,716) | 20.4% (23,620) | 23.6% (11,096) | 21.1% (32,376) | 24.0% (2,340) |
| Often | 2.8% (4,641) | 2.0% (2,281) | 5.0% (2,360) | 2.7% (4,183) | 4.7% (458) |
| All of the time | 0.0% (1) | 0.0% (0) | 0.0% (1) | 0.0% (1) | 0.0% (0) |
| Nap during the day |  |  |  |  |  |
| Never/rarely | 55.8% (91,166) | 56.8% (66,032) | 53.2% (25,134) | 56.1% (86,168) | 50.9% (4,998) |
| Sometimes | 38.9% (63,559) | 38.5% (44,713) | 39.9% (18,846) | 38.7% (59,441) | 42.0% (4,118) |
| Usually | 5.3% (8,728) | 4.7% (5,439) | 7.0% (3,289) | 5.2% (8,033) | 7.1% (695) |
| How easy find getting up in morning |  |  |  |  |  |
| Not at all easy | 4.0% (6,556) | 2.8% (3,234) | 7.0% (3,322) | 3.8% (5,818) | 7.5% (738) |
| Not very easy | 13.7% (22,430) | 12.0% (13,969) | 17.9% (8,461) | 13.5% (20,655) | 18.1% (1,775) |
| Fairly easy | 49.2% (80,446) | 50.9% (59,167) | 45.1% (21,279) | 49.5% (75,987) | 45.5% (4,459) |
| Very easy | 33.0% (53,930) | 34.3% (39,795) | 29.9% (14,135) | 33.3% (51,101) | 28.9% (2,829) |
| Job involves night shift work |  |  |  |  |  |
| Never/rarely | 95.0% (155,418) | 94.7% (110,170) | 95.6% (45,248) | 94.9% (146,032) | 95.5% (9,386) |
| Sometimes | 2.8% (4,573) | 3.0% (3,437) | 2.4% (1,136) | 2.8% (4,321) | 2.6% (252) |
| Usually | 0.8% (1,320) | 0.9% (1,021) | 0.6% (299) | 0.8% (1,257) | 0.6% (63) |
| Always | 1.4% (2,331) | 1.5% (1,705) | 1.3% (626) | 1.4% (2,207) | 1.3% (124) |
| Metabolic Equivalent TasK (MET) minutes per week quartiles |  |  |  |  |  |
| Q1 (0-813) | 25.0% (32,985) | 24.0% (22,771) | 27.6% (10,214) | 24.8% (30,802) | 28.5% (2,183) |
| Q2 (815-1815) | 25.0% (32,907) | 25.3% (23,992) | 24.1% (8,915) | 25.0% (31,002) | 24.8% (1,905) |
| Q3 (1816.8-3679) | 25.0% (32,900) | 25.4% (24,079) | 23.9% (8,821) | 25.1% (31,146) | 22.9% (1,754) |
| Q4 (3679.2-19278) | 25.0% (32,919) | 25.2% (23,890) | 24.4% (9,029) | 25.1% (31,090) | 23.8% (1,829) |
| Coffee intake |  |  |  |  |  |
| 0-1 cups/day | 48.9% (79,880) | 48.6% (56,417) | 49.7% (23,463) | 48.8% (74,916) | 50.6% (4,964) |
| 2-3 cups/day | 30.9% (50,434) | 31.2% (36,259) | 30.0% (14,175) | 30.9% (47,495) | 30.0% (2,939) |
| 4-5 cups/day | 13.7% (22,378) | 13.8% (16,071) | 13.4% (6,307) | 13.8% (21,115) | 12.9% (1,263) |
| 6 or more cups/day | 6.5% (10,645) | 6.3% (7,373) | 6.9% (3,272) | 6.5% (10,007) | 6.5% (638) |
| Tea intake |  |  |  |  |  |
| 0-2 cups/day | 39.9% (65,146) | 39.9% (46,322) | 39.9% (18,824) | 39.9% (61,334) | 38.9% (3,812) |
| 3-5 cups/day | 40.7% (66,509) | 41.2% (47,875) | 39.5% (18,634) | 40.7% (62,552) | 40.4% (3,957) |
| 6-8 cups/day | 15.5% (25,247) | 15.1% (17,582) | 16.2% (7,665) | 15.4% (23,677) | 16.0% (1,570) |
| 9 or more cups/day | 4.0% (6,458) | 3.8% (4,382) | 4.4% (2,076) | 3.9% (5,997) | 4.7% (461) |
| BMI |  |  |  |  |  |
| Underweight | 0.5% (805) | 0.5% (554) | 0.5% (251) | 0.5% (752) | 0.5% (53) |
| Healthy weight | 31.8% (51,756) | 32.6% (37,725) | 29.8% (14,031) | 32.0% (49,018) | 28.1% (2,738) |
| Overweight | 42.7% (69,517) | 43.4% (50,215) | 41.0% (19,302) | 42.9% (65,697) | 39.1% (3,820) |
| Obese | 25.0% (40,748) | 23.6% (27,286) | 28.6% (13,462) | 24.6% (37,600) | 32.3% (3,148) |
| Takes risks | 26.8% (42,123) | 27.2% (30,430) | 25.7% (11,693) | 26.7% (39,528) | 27.6% (2,595) |
| Smoking status |  |  |  |  |  |
| Never | 54.8% (89,408) | 56.1% (65,087) | 51.6% (24,321) | 55.1% (84,450) | 50.7% (4,958) |
| Previous | 35.0% (57,079) | 34.0% (39,390) | 37.5% (17,689) | 34.9% (53,463) | 37.0% (3,616) |
| Current | 10.2% (16,646) | 9.9% (11,511) | 10.9% (5,135) | 10.1% (15,449) | 12.3% (1,197) |
| Alcohol intake frequency |  |  |  |  |  |
| Daily/almost daily | 20.0% (32,726) | 19.9% (23,191) | 20.2% (9,535) | 20.1% (30,891) | 18.7% (1,835) |
| 3-4 times a week | 23.1% (37,800) | 23.9% (27,791) | 21.2% (10,009) | 23.3% (35,787) | 20.5% (2,013) |
| Once or twice a week | 26.0% (42,560) | 26.4% (30,738) | 25.0% (11,822) | 26.1% (40,153) | 24.5% (2,407) |
| 1-3 times a month | 11.2% (18,364) | 11.1% (12,865) | 11.6% (5,499) | 11.2% (17,199) | 11.9% (1,165) |
| Special occasions only | 11.4% (18,594) | 10.8% (12,547) | 12.8% (6,047) | 11.2% (17,225) | 14.0% (1,369) |
| Never | 8.3% (13,558) | 7.9% (9,174) | 9.3% (4,384) | 8.2% (12,536) | 10.4% (1,022) |
| Have had menopause (women only) | 76.1% (64,905) | 72.4% (41,509) | 83.6% (23,396) | 75.7% (60,234) | 82.2% (4,671) |
| Frequency depressed mood past 2 weeks category |  |  |  |  |  |
| Not at all | 76.1% (118,835) | 80.5% (89,626) | 65.3% (29,209) | 76.8% (112,731) | 66.0% (6,104) |
| Several days | 18.5% (28,886) | 15.7% (17,518) | 25.4% (11,368) | 18.1% (26,596) | 24.8% (2,290) |
| More than half the days | 3.3% (5,098) | 2.5% (2,830) | 5.1% (2,268) | 3.2% (4,633) | 5.0% (465) |
| Nearly every day | 2.1% (3,253) | 1.2% (1,345) | 4.3% (1,908) | 1.9% (2,862) | 4.2% (391) |
| Are a worrier | 56.4% (89,744) | 51.6% (58,247) | 68.2% (31,497) | 55.7% (83,381) | 66.4% (6,363) |
| Overall health rating |  |  |  |  |  |
| Excellent | 14.9% (24,332) | 16.9% (19,584) | 10.1% (4,748) | 15.3% (23,509) | 8.4% (823) |
| Good | 58.2% (94,846) | 60.7% (70,349) | 52.1% (24,497) | 58.7% (89,952) | 50.0% (4,894) |
| Fair | 22.0% (35,870) | 19.3% (22,434) | 28.6% (13,436) | 21.5% (32,954) | 29.8% (2,916) |
| Poor | 4.9% (7,927) | 3.1% (3,581) | 9.2% (4,346) | 4.4% (6,778) | 11.7% (1,149) |
