## Supplementary material for "Insomnia symptom prevalence in England: A comparison of self-reported data and primary care records in the UK Biobank": Table S4 Stratification Table

TABLE S4 Self-reported and primary care insomnia symptom cases stratified by sociodemographics, lifestyle, sleep & health characteristics

|  | **Self-reported insomnia case** | **Primary care insomnia case** |
| --- | --- | --- |
| Row % |  |  |
| Sex |  |  |
| Female | 32.8 | 6.7 |
| Male | 24.2 | 5.2 |
| Age |  |  |
| Under 45 | 20.8 | 4.5 |
| 45-54 | 26.9 | 5.5 |
| 55-64 | 30.9 | 6.4 |
| 65 or over | 31.6 | 6.6 |
| Ethnic group |  |  |
| White | 29.2 | 6.0 |
| Mixed | 30.8 | 6.2 |
| Asian/Asian British | 22.0 | 6.2 |
| Black/Black British | 20.7 | 6.0 |
| Chinese | 19.1 | 5.1 |
| Other | 24.7 | 7.4 |
| Average household income (before tax) |  |  |
| <£18,000 | 34.7 | 7.4 |
| £18,000-£30,999 | 29.1 | 6.1 |
| £31,000-£51,999 | 26.2 | 5.3 |
| £52,000-£100,000 | 24.4 | 4.8 |
| >£100,000 | 21.2 | 4.0 |
| Index of Multiple Deprivation for England Quartiles |  |  |
| Q1 (0.76-7.85) | 27.2 | 5.7 |
| Q2 (7.86-13.59) | 28.0 | 5.8 |
| Q3 (13.6-23.85) | 28.9 | 5.9 |
| Q4 (23.86-81.59) | 31.6 | 6.8 |
| Current employment status |  |  |
| Paid employment / self-employed | 25.3 | 5.1 |
| Retired | 32.3 | 6.7 |
| Other | 38.4 | 9.1 |
| Highest qualification |  |  |
| None | 34.9 | 7.3 |
| College/University degree | 25.2 | 5.3 |
| A/AS levels or equivalent | 26.8 | 5.6 |
| O levels/GCSEs or equivalent | 29.8 | 6.1 |
| CSEs or equivalent | 29.5 | 5.9 |
| NVQ/HND/HNC or equivalent | 29.1 | 5.8 |
| Other professional qualifications | 29.4 | 6.5 |
| Household size |  |  |
| 1 person | 32.0 | 7.0 |
| 2 people | 30.3 | 6.1 |
| 3-5 people | 25.1 | 5.2 |
| 6 or more people | 23.7 | 5.4 |
| Live with spouse/partner |  |  |
| No | 29.7 | 7.0 |
| Yes | 28.0 | 5.6 |
| Home area population density |  |  |
| Urban | 29.1 | 6.0 |
| Town | 29.1 | 6.6 |
| Rural | 26.7 | 5.7 |
| Sleep duration |  |  |
| 3-4 hours | 85.5 | 20.0 |
| 5-6 hours | 49.5 | 8.7 |
| 7-8 hours | 21.4 | 4.8 |
| 9 or more hours | 21.6 | 5.8 |
| Chronotype |  |  |
| Definite morning | 29.2 | 6.3 |
| Morning more than evening | 28.2 | 5.5 |
| No preference | 27.9 | 5.8 |
| Evening more than morning | 28.6 | 6.1 |
| Definite evening | 33.3 | 6.9 |
| Snore |  |  |
| No | 29.3 | 5.7 |
| Yes | 27.2 | 6.2 |
| Doze/fall asleep during the day when don’t mean to |  |  |
| Never/rarely | 27.2 | 5.6 |
| Sometimes | 32.0 | 6.7 |
| Often | 50.9 | 9.9 |
| All of the time | 100.0 | 0.0 |
| Nap during the day |  |  |
| Never/rarely | 27.6 | 5.5 |
| Sometimes | 29.7 | 6.5 |
| Usually | 37.7 | 8.0 |
| How easy find getting up in morning |  |  |
| Not at all easy | 50.7 | 11.3 |
| Not very easy | 37.7 | 7.9 |
| Fairly easy | 26.5 | 5.5 |
| Very easy | 26.2 | 5.2 |
| Job involves night shift work |  |  |
| Never/rarely | 29.1 | 6.0 |
| Sometimes | 24.8 | 5.5 |
| Usually | 22.7 | 4.8 |
| Always | 26.9 | 5.3 |
| Metabolic Equivalent TasK (MET) minutes per week quartiles |  |  |
| Q1 (0-813) | 31.0 | 6.6 |
| Q2 (815-1815) | 27.1 | 5.8 |
| Q3 (1816.8-3679) | 26.8 | 5.3 |
| Q4 (3679.2-19278) | 27.4 | 5.6 |
| Coffee intake |  |  |
| 0-1 cups/day | 29.4 | 6.2 |
| 2-3 cups/day | 28.1 | 5.8 |
| 4-5 cups/day | 28.2 | 5.6 |
| 6 or more cups/day | 30.7 | 6.0 |
| Tea intake |  |  |
| 0-2 cups/day | 28.9 | 5.9 |
| 3-5 cups/day | 28.0 | 5.9 |
| 6-8 cups/day | 30.4 | 6.2 |
| 9 or more cups/day | 32.1 | 7.1 |
| BMI |  |  |
| Underweight | 31.2 | 6.6 |
| Healthy weight | 27.1 | 5.3 |
| Overweight | 27.8 | 5.5 |
| Obese | 33.0 | 7.7 |
| Takes risks |  |  |
| No | 29.3 | 5.9 |
| Yes | 27.8 | 6.2 |
| Smoking status |  |  |
| Never | 27.2 | 5.5 |
| Previous | 31.0 | 6.3 |
| Current | 30.8 | 7.2 |
| Alcohol intake frequency |  |  |
| Daily/almost daily | 29.1 | 5.6 |
| 3-4 times a week | 26.5 | 5.3 |
| Once or twice a week | 27.8 | 5.7 |
| 1-3 times a month | 29.9 | 6.3 |
| Special occasions only | 32.5 | 7.4 |
| Never | 32.3 | 7.5 |
| Have had menopause (women only) |  |  |
| No | 22.5 | 5.0 |
| Yes | 36.0 | 7.2 |
| Frequency depressed mood past 2 weeks |  |  |
| Not at all | 24.6 | 5.1 |
| Several days | 39.4 | 7.9 |
| More than half the days | 44.5 | 9.1 |
| Nearly every day | 58.7 | 12.0 |
| Are a worrier |  |  |
| No | 21.2 | 4.6 |
| Yes | 35.1 | 7.1 |
| Overall health rating |  |  |
| Excellent | 19.5 | 3.4 |
| Good | 25.8 | 5.2 |
| Fair | 37.5 | 8.1 |
| Poor | 54.8 | 14.5 |
