## Supplementary material for "Insomnia symptom prevalence in England: A comparison of self-reported data and primary care records in the UK Biobank": Tables S5-10 Sensitivity Analyses

**Tables S5-S10 Sensitivity Analyses Cross-Tabulations**

**TABLE S5** Cross-tabulation of Self-Reported Insomnia Symptom Cases and Primary Care Insomnia Symptom Cases (defined as having an insomnia Read code in the 12 months prior to baseline).

| **Primary care insomnia symptom case** | | **Self-reported insomnia symptom case** | | |
| --- | --- | --- | --- | --- |
|  |  | No | Yes | Total |
| No | Frequency | 115,570 | 46,269 | 161,839 |
|  | Row % | 71.4 | 28.6 | 100.0 |
|  | Column % | 99.3 | 97.8 | 98.8 |
| Yes | Frequency | 844 | 1,065 | 1,909 |
|  | Row % | 44.2 | 55.8 | 100.0 |
|  | Column % | 0.7 | 2.3 | 1.2 |
| Total | Frequency | 116,414 | 47,334 | 163,748 |
|  | Row % | 71.1 | 28.9 | 100.0 |
|  | Column % | 100.0 | 100.0 | 100.0 |

**TABLE S6** Cross-tabulation of Self-Reported Insomnia Symptom Cases and Primary Care Insomnia Symptom Cases (defined as having an insomnia Read code in the four weeks prior to baseline).

| **Primary care insomnia symptom case** | | **Self-reported insomnia symptom case** | | |
| --- | --- | --- | --- | --- |
|  |  | No | Yes | Total |
| No | Frequency | 116,342 | 47,208 | 163,550 |
|  | Row % | 71.1 | 28.9 | 100.0 |
|  | Column % | 99.9 | 99.7 | 99.9 |
| Yes | Frequency | 72 | 126 | 198 |
|  | Row % | 36.4 | 63.6 | 100.0 |
|  | Column % | 0.1 | 0.3 | 0.1 |
| Total | Frequency | 116,414 | 47,334 | 163,748 |
|  | Row % | 71.1 | 28.9 | 100.0 |
|  | Column % | 100.0 | 100.0 | 100.0 |

**TABLE S7** Cross-tabulation of Self-Reported Insomnia Symptom Cases and Primary Care Insomnia Symptom Cases (defined as having an insomnia Read code prior to baseline and a prescription for a hypnotic within 90 days of the Read code).

| **Primary care insomnia symptom case** | | **Self-reported insomnia symptom case** | | |
| --- | --- | --- | --- | --- |
|  |  | No | Yes | Total |
| No | Frequency | 115,134 | 45,591 | 160,725 |
|  | Row % | 71.6 | 28.4 | 100.0 |
|  | Column % | 98.9 | 96.3 | 98.2 |
| Yes | Frequency | 1,280 | 1,743 | 3,023 |
|  | Row % | 42.3 | 57.7 | 100.0 |
|  | Column % | 1.1 | 3.7 | 1.9 |
| Total | Frequency | 116,414 | 47,334 | 163,748 |
|  | Row % | 71.1 | 28.9 | 100.0 |
|  | Column % | 100.0 | 100.0 | 100.0 |

**TABLE S8** Cross-tabulation of Self-Reported Insomnia Symptom Cases and Primary Care Insomnia Symptom Cases (defined as having an insomnia Read code in the 12 months prior to baseline and a prescription for a hypnotic within 90 days of the Read code).

| **Primary care insomnia symptom case** | | **Self-reported insomnia symptom case** | | |
| --- | --- | --- | --- | --- |
|  |  | No | Yes | Total |
| No | Frequency | 116,224 | 46,981 | 163,205 |
|  | Row % | 71.2 | 28.8 | 100.0 |
|  | Column % | 99.8 | 99.3 | 99.7 |
| Yes | Frequency | 190 | 353 | 543 |
|  | Row % | 35.0 | 65.0 | 100.0 |
|  | Column % | 0.2 | 0.8 | 0.3 |
| Total | Frequency | 116,414 | 47,334 | 163,748 |
|  | Row % | 71.1 | 28.9 | 100.0 |
|  | Column % | 100.0 | 100.0 | 100.0 |

**TABLE S9** Cross-tabulation of Self-Reported Insomnia Symptom Cases and Primary Care Insomnia Symptom Cases (defined as having an insomnia Read code in the four weeks prior to baseline and a prescription for a hypnotic within 90 days of the Read code).

| **Primary care insomnia symptom case** | | **Self-reported insomnia symptom case** | | |
| --- | --- | --- | --- | --- |
|  |  | No | Yes | Total |
| No | Frequency | 116,400 | 47,297 | 163,697 |
|  | Row % | 71.1 | 28.9 | 100.0 |
|  | Column % | 99.99 | 99.92 | 99.97 |
| Yes | Frequency | 14 | 37 | 51 |
|  | Row % | 27.5 | 72.6 | 100.0 |
|  | Column % | 0.01 | 0.08 | 0.03 |
| Total | Frequency | 116,414 | 47,334 | 163,748 |
|  | Row % | 71.1 | 28.9 | 100.0 |
|  | Column % | 100.0 | 100.0 | 100.0 |

**TABLE S10** Cross-tabulation of Self-Reported Insomnia Symptom Cases and Primary Care Insomnia Symptom Cases (defined as having a hypnotic prescription prior to baseline).

| **Primary care insomnia symptom case** | | **Self-reported insomnia symptom case** | | |
| --- | --- | --- | --- | --- |
|  |  | No | Yes | Total |
| No | Frequency | 105,940 | 39,249 | 145,189 |
|  | Row % | 73.0 | 27.0 | 100.0 |
|  | Column % | 91.00 | 82.9 | 88.7 |
| Yes | Frequency | 10,474 | 8,085 | 18,559 |
|  | Row % | 56.4 | 43.6 | 100.0 |
|  | Column % | 9.00 | 17.1 | 11.3 |
| Total | Frequency | 116,414 | 47,334 | 163,748 |
|  | Row % | 71.1 | 28.9 | 100.0 |
|  | Column % | 100.0 | 100.0 | 100.0 |
